# Automated Interpretable Discovery of Heterogeneous Treatment Effectiveness: A Covid-19 Case Study

**DOI:** 10.1101/2021.10.30.21265430

**Authors:** Benjamin J. Lengerich, Mark E. Nunally, Yin Aphinyanaphongs, Rich Caruana

## Abstract

Testing multiple treatments for heterogeneous (varying) effectiveness with respect to many underlying risk factors requires many pairwise tests; we would like to instead automatically discover and visualize patient archetypes and predictors of treatment effectiveness using multitask machine learning. In this paper, we present a method to estimate these heterogeneous treatment effects with an interpretable hierarchical framework that uses additive models to visualize expected treatment benefits as a function of patient factors (identifying personalized treatment benefits) and concurrent treatments (identifying combinatorial treatment benefits). This method achieves state-of-the-art predictive power for Covid-19 in-hospital mortality and interpretable identification of heterogeneous treatment benefits. We first validate this method on the large public MIMIC-IV dataset of ICU patients to test recovery of heterogeneous treatment effects. Next we apply this method to a proprietary dataset of over 3000 patients hospitalized for Covid-19, and find evidence of heterogeneous treatment effectiveness predicted largely by indicators of inflammation and thrombosis risk: patients with few indicators of thrombosis risk benefit most from treatments against inflammation, while patients with few indicators of inflammation risk benefit most from treatments against thrombosis. This approach provides an automated methodology to discover heterogeneous and individualized effectiveness of treatments.

## 1 Introduction

Medical treatments can have different effectiveness for different patients based on a wide variety of factors including patient histories, comorbidities, and concurrent treatments; for this reason, a wide variety of statistical tools [1–4] have been developed to robustly estimate heterogeneous treatment effects (HTE) and individualized treatment effects (ITE) as functions of patient features. However, independently testing multiple treatments for heterogeneous effects with respect to many underlying patient risk factors requires large numbers of pairwise tests. To avoid reducing statistical power, we would like an automated method which uses multi-task learning to simultaneously discover heterogeneous treatments effects of many treatments and risk factors.

In this paper, we focus on the S-learner strategy [5] of estimating the conditional average treatment effect (CATE). This strategy first seeks 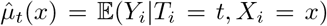 using a single model, where *X*_*i*_ are patient factors and *T*_*i*_ are treatments; the heterogeneous treatment effects are then provided as 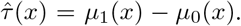. While this strategy is straightforward, there are statistical challenges with modeling interactions between treatments and risk factors. To overcome this challenge, we propose to use multitask learning to share statistical power between treatments to identify patient representations which predict the effectiveness of several related medications. This can be interpreted as an S-learner where *T*_*i*_ is a vector of treatments for patient *i* rather than a scalar indicator of a single treatment.

Our proposed framework (Figure 1) uses additive models to visualize expected treatment benefits as a function of patient factors (identifying personalized treatment benefits) and concurrent treatments (identifying interactive treatment benefits). This framework has three main benefits: (1) it is a multi-task learning method which shares statistical power between related treatments, (2) it inherits the interpretability of additive models, allowing us to visualize impact of patient risk factors, and (3) it naturally extends to continuous-valued, dosed treatments. This compares against prior works which select linear regression models based on subgroup identification [6–8], and are thus limited to clustering patients, or regression trees [9, 10] which do not provide interpretable maps linking risk factors to treatment benefits.

**Figure 1:**
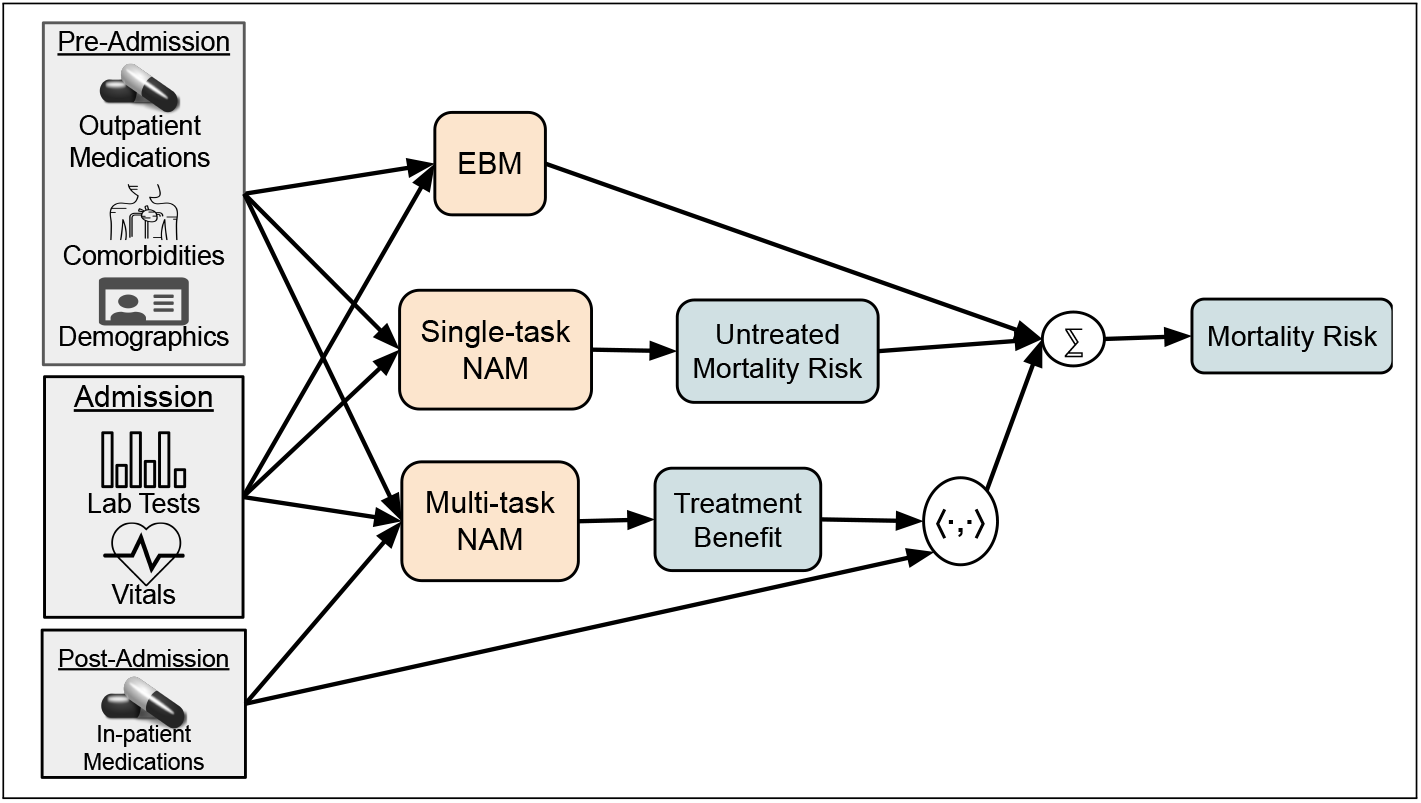
Architecture to estimate latent personalized treatment benefits. Gray boxes indicate observed data, orange boxes are learned models, and blue boxes are latent variables. We first train an Explainable Boosting Machine (EBM) to predict mortality risk from pre-treatment features to generate baseline mortality risk. This model, based on gradient-boosted trees, captures discontinuities in risk, but is not differentiable and so must be trained in isolation from the rest of the architecture. After the EBM is trained, we train the single-task Neural Additive Model (NAM) and the multi-task NAM in parallel to decompose mortality risk into untreated mortality risk and personalized treatment benefits. This framework provides state-of-art mortality risk prediction and decomposes the risk into interpretable additive factors contributing to underlying risk and treatment benefits.

In this paper, we first validate the proposed method on the MIMIC-IV dataset to test its recovery of HTEs. Next we apply this method to a dataset of over 3000 patients hospitalized for Covid-19, and find evidence of HTEs predicted largely by indicators of inflammation and thrombosis risk: patients with few indicators of thrombosis risk benefit most from treatments against inflammation, while patients with few indicators of inflammation risk benefit most from treatments against thrombosis. This approach provides an automated methodology to discover heterogeneous and individualized effectiveness of treatments, which can be followed up by targeted statistical tests and clinical studies.

## 2 Materials and Methods

### Cohort

Our dataset consists of 11080 total hospitalized patients who had lab-confirmed cases of Covid-19 from March 2020 to August 2020. To filter out patients who were hospitalized for reasons other than Covid-19, we excluded patients who have indicators of (1) pregnancy: outpatient prenatal vitamins, in-patient oxytocics, folic acid preparations; or (2) scheduled surgery: urinary tract radiopaque diagnostics, laxatives, general anesthetics, antiemetic/antivertigo agents, or antiparasitics. We also require that the patients have recorded temperature, age, BMI, and Admission Day. Finally, we remove patients who died within six hours of admission.

To correct for patient risk confounding, we include pre-admission features (demographics, comorbidities, and outpatient medications), and features taken on admission (vitals and initial in-patient lab tests). We exclude any measurement taken within 24 hours of the patient mortality. The full list of features and more details regarding the cohort are provided in Sec. S1.

### Treatment and Outcome Measure Construction

To ensure a proper linking of lab values, treatments, and outcomes, we consider only treatments that are administered within 24 hours of the initial lab test and at least 24 hours before mortality. We do not consider dosage of treatments in this study. Our outcome is in-hospital mortality. The cohort has an average mortality rate of 18.1%; as shown in Figure S2a, the mortality rate peaked over 25% in March 2020 and decreased to less than 5% in August 2020.

### 2.1 Methods

Our goal in this study is to decompose the patient mortality risk into underlying risk factors and treatment benefits:

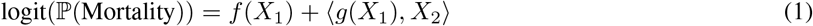

where *X*_1_ are features observed at or prior to hospital admission, and *X*_2_ are in-patient treatments. The function *g*(*X*_1_) estimates the expected benefit of administering treatments for each patient; the inner product ⟨*g*(*X*_1_), *X*_2_⟩ converts this potential benefit into an estimated scalar benefit under the observed treatments *X*_2_. This model decomposes patient mortality risk into a background risk and a treatment benefit. There are two main challenges in estimation: estimating *f* the background mortality risk model and estimating *g* the personalized treatment benefit model. In this paper, we use generalized additive models for both *f* and *g*. `Python` code for this model is available at github.com/blengerich/ContextualizedGAM.

### Background Mortality Risk Model

We use generalized additive models (GAMs) to model patient mortality risk. GAMs are a version of logistic regression that are able to model non-linear effects [11]. While logistic regression summarizes the influence of each feature with a single coefficient, GAMs estimate the influence of a feature for every value the feature can take on as a graph. This means that GAMs naturally accommodate non-linear effects, which improves both model accuracy and interpretation. Non-linear effects are particularly important when features have multiple regions of high or low risk (e.g., both hyperthermia and hypothermia are associated with high risk). We use tree-based GAMs [12] implemented in the Python `Interpret` package^1^. These “Explainable Boosting Machine” (EBM) GAMs are invariant to all monotonic feature transforms, so log-transforms of lab values are not necessary.

### Estimating Personalized Treatment Benefits

To estimate *g*(*X*_1_, *X*_2_), we use a neural additive model (NAM) [13]. This provides an interpretable model which relates patient risk factors to expected treatment benefits. We also allow *g* (the estimated treatment benefits) to vary with *X*_2_ (in-patient medications) to identify combination therapies which have interaction effects. By estimating a multivariate *g*, we share power between multiple “tasks” of treatments and improve estimation of patient representations and treatment responses.

Finally, while the tree-based EBM of background mortality risk (*f*) is highly accurate (due to its capacity to fit discontinuities of treatment protocols), it is not differentiable and thus must be pre-trained and frozen before training *g*. For this reason, we estimate a third function *h*(*X*_1_) as a NAM to adjust the EBM *f* by removing effects which were initially estimated as effects in *f* but are better captured as effects in *g*. This model architecture is shown in Figure 1.

### Method Validation with MIMIC-IV

Before applying this method for estimation of heterogeneous treatment effectiveness to the novel Covid-19 disease, we first validate recovery of known biomedical pathways from observational data. To do this, we use the MIMIC-IV dataset, which consists of 53,150 patient admitted to an ICU, and records detailed lab values, treatments, and outcomes for all patients. Results of this analysis are shown in Section S4, and demonstrate that this methodology identifies known and plausible heterogeneous treatment effectiveness, based on patient age and hemoglobin levels, from observational data.

## 3 Results

Our approach decomposes mortality risk into several factors: underlying risk, homogeneous treatment effects, and heterogeneous treatment effects.

### 3.1 Risk Factors

The risk model is accurate, achieving an ROC of 0.912 ± 0.001 and an F1-score of 0.598 ± 0.002 on held-out patients, outperforming a logistic regression model which achieves an ROC of 0.859 ± 0.001 and F1-score of 0.455 ± 0.002 (confidence intervals estimated by bootstrap resampling). The most important features to the background risk model are (in decreasing order): Neutrophil-Lymphocyte Ratio (NLR), Temperature, Age, Procalcitonin and C-Reactive Protein. While elevated NLR is a strong predictor of mortality, none of these risk factors solely determine mortality (Figure S1 shows that many patients with low NLR levels nevertheless have large probabilities of mortality).

#### Comorbidities and Pre-Admission Medications

The baseline mortality risks associated with comorbidities and pre-admission out-patient medications are shown in Figure 2. In general, these effects of commodities and out-patient medications concord with mechanisms of mortality involving inflammation and/or thromboses. The most protective association is valve replacement, for which patients are typically prescribed long-term anti-coagulants. The secondmost protective association is platelet aggregation inhibitors (low-dose aspirin), which is an anti-coagulant. The most deleterious associations are congestive heart failure, hypertension, and myocardial infarction.

**Figure 2:**
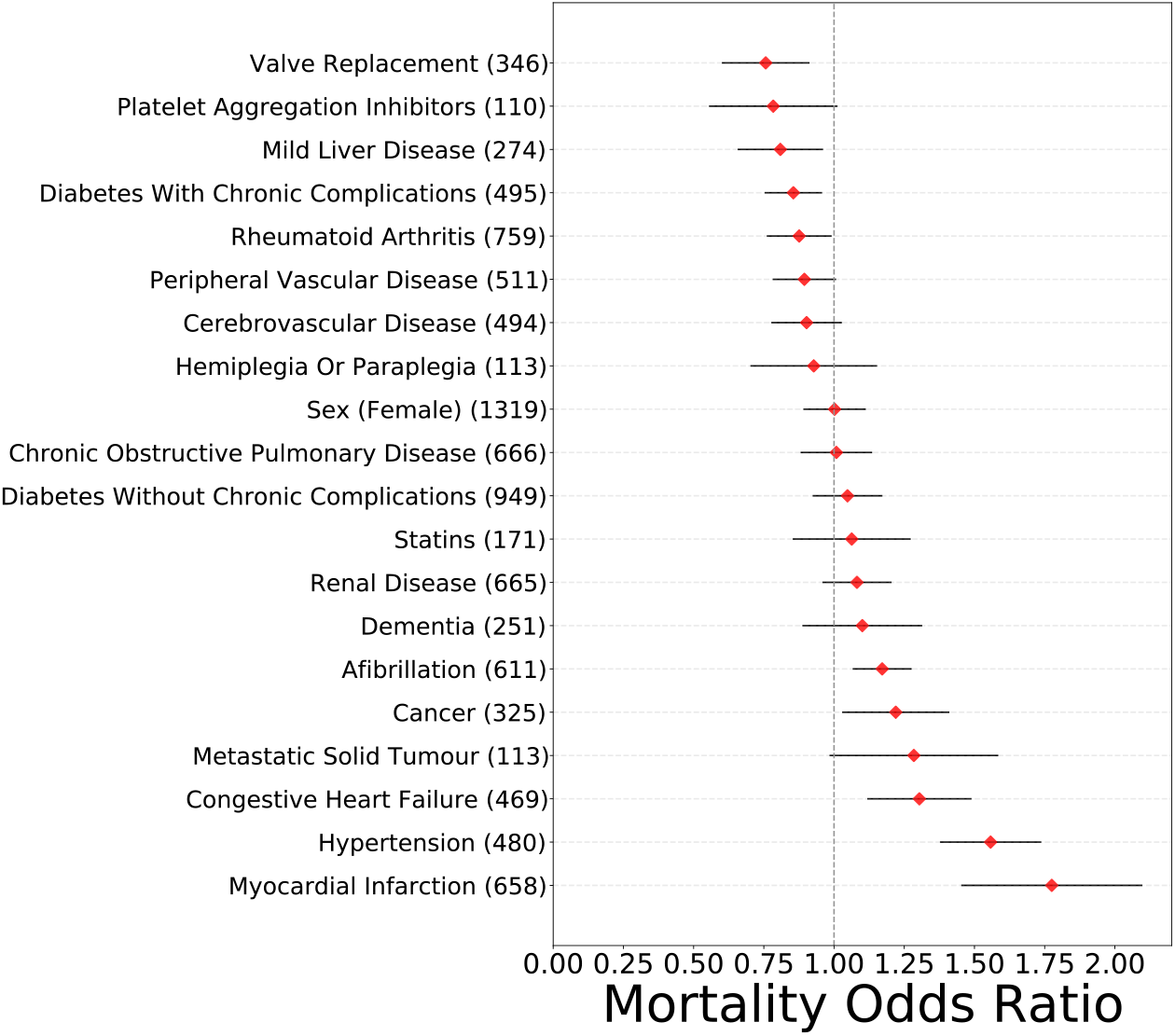
Mortality risk conferred by pre-admission comorbidities and medications, with 95% confidence intervals. For each treatment, the number in parentheses indicates the sample size.

#### Demographics and Vital Signs

As shown in Figure 3, patient age and initial temperature on admission are strong predictors of mortality of hospitalized patients, while patient body mass index (BMI) is not a strong risk factor (after any selection bias in admission protocols).

**Figure 3:**
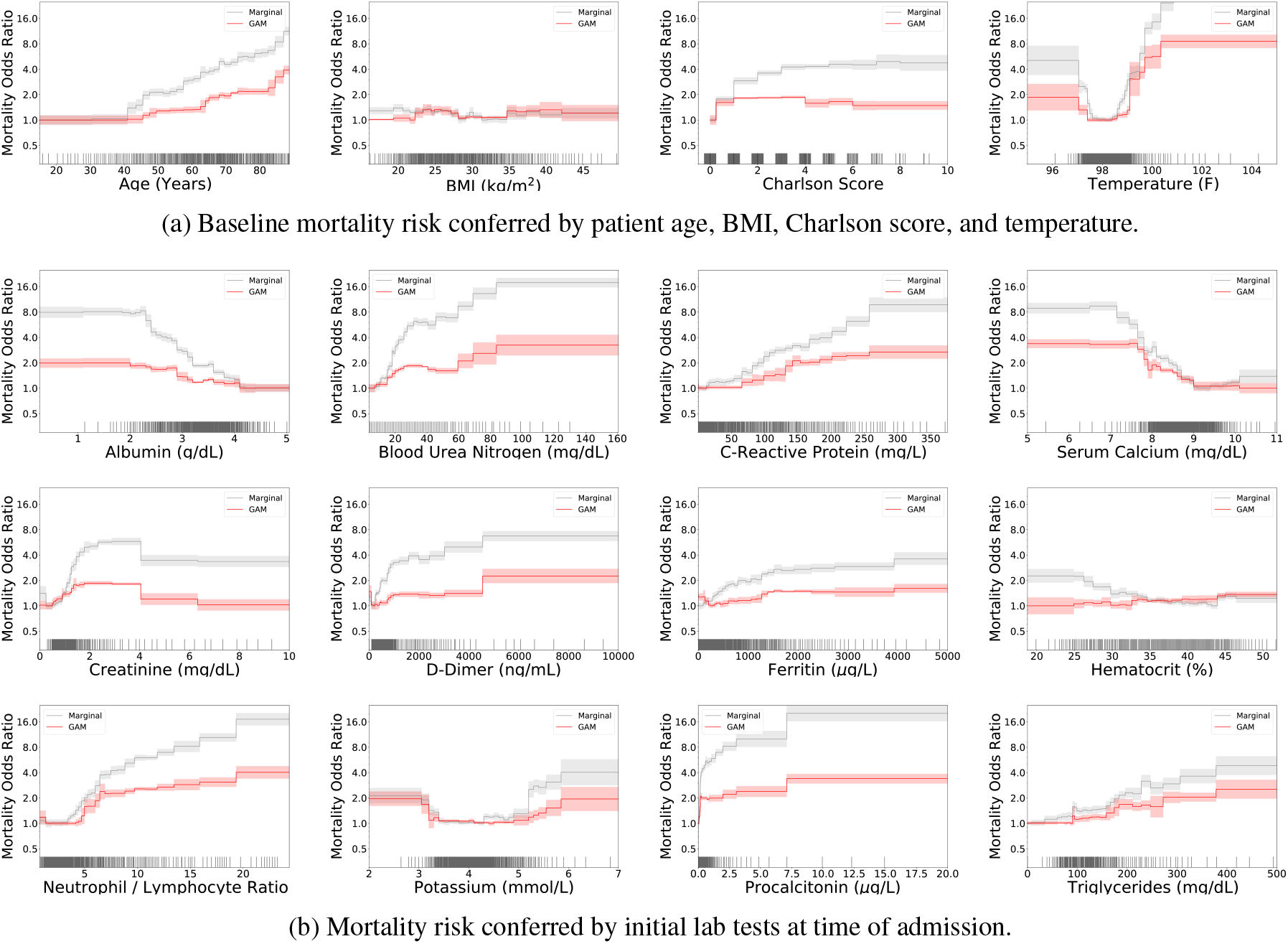
Mortality risk conferred by demographics, vitals, and lab tests. In each pane, we show both the effect estimated by univariable marginalization (gray), and the multivariable GAM which corrects for other risk factors (red). Each black tick mark along the horizontal indicates 10 patients, with noise added to visualize data density.

#### Lab Tests

The effects of 12 lab tests are shown in Figure 3. The largest effect is for Neutrophil / Lymphocyte Ratio (NLR), a measure of inflammation and an indicator of Covid-19 severity [14, 15]. Procalcitonin and C-Reactive Protein also increase likelihood of mortality. Also of note are serum calcium, for which no elevated level is associated with increased risk, and Creatinine, which shows a step-function drop in risk at 4 mg/dL, a common threshold for dialysis decisions. Finally, Hematocrit exhibits opposite effects when estimated in isolation or after correcting for other factors – marginalization associates decreased Hematocrit with increased mortality risk, while the multivariable GAM identifies increased Hematocrit with increased mortality risk.

### 3.2 Homogeneous Treatment Effects

For homogeneous average treatment effects (ATEs), we are interested in excess mortality expressed as the adjusted risk difference (ARD) Σ*i Y*_*i*_*T*_*i*_ −Σ_i_ ℙ(mortality|*X*_*i*_), where *T*_*i*_ is the treatment indicator for patient *i* and 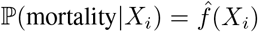 is given by the background mortality risk model. For both measures, smaller values indicate protective associations while larger values indicate harm (a treatment with no association would have ARD=0 and ARR=1).

Figure 4 shows the mean mortality risk conferred by in-patient medications (without accounting for HTEs) after correcting for all other observed risk factors. The vast majority of these treatments do not show statistical evidence of homogeneous treatment benefit. There is some evidence of a benefit of thyroid hormones, concording with observed dysregulation of thyroid hormones in Covid-19 patients [16]. In addition, there is some evidence of a protective effect of NSAIDs (defined as either Ibuprofen or Ketorolac) in a small sample size of 101 treated patients. We also see a possibility of a beneficial effect of direct factor Xa inhibitors; however, the confidence intervals are very wide and it is difficult to distinguish any protective effect from association with administration prior to patient discharge.

**Figure 4:**
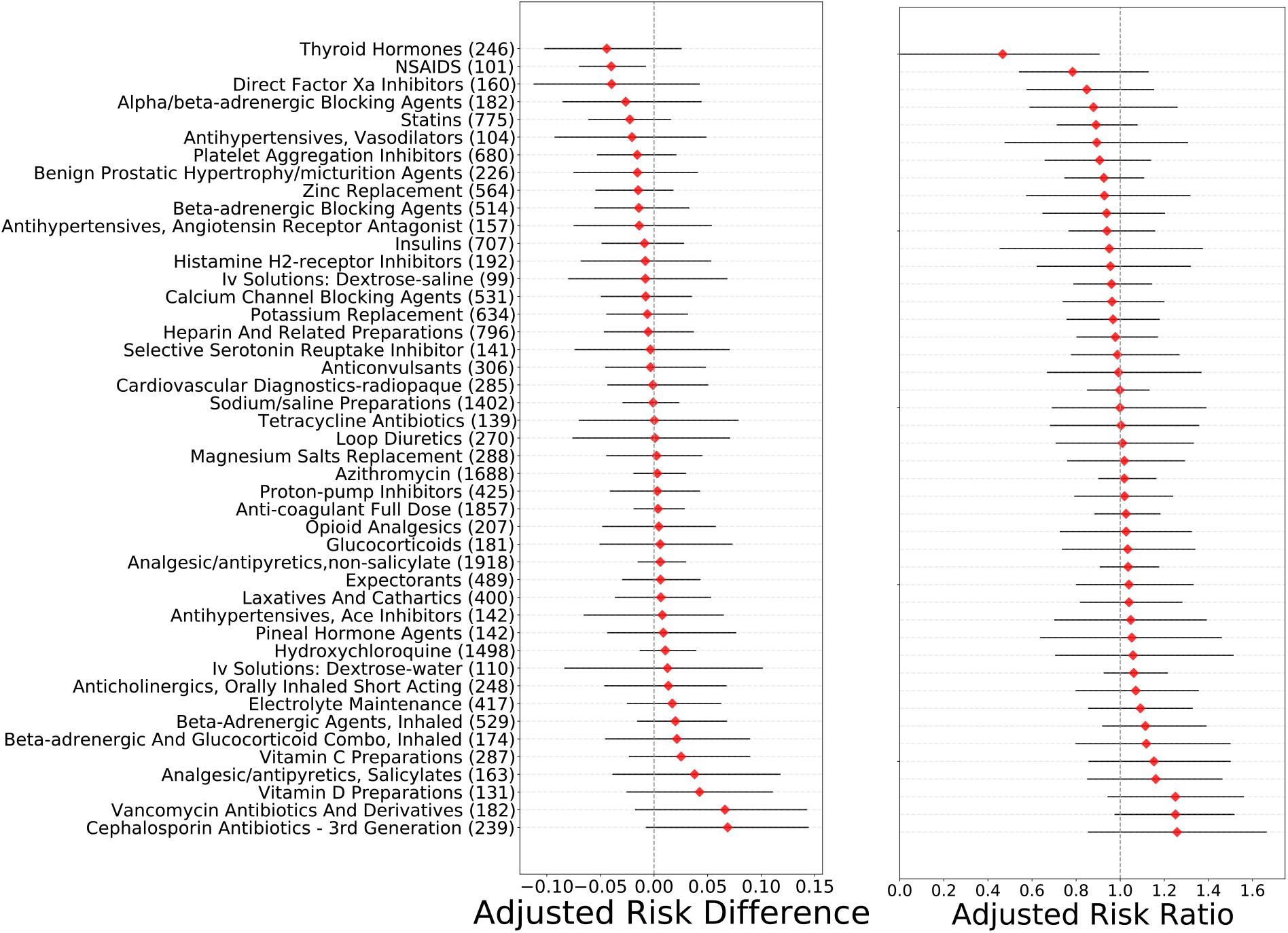
Homogeneous treatment effects of in-patient medications, calculated as the adjusted risk difference (left) and the adjusted risk ratio (right). For each treatment, the number in parentheses is the number of patients treated with this medication in the dataset.

### 3.3 Heterogeneous Treatment Effects

Next, we turn to the HTEs of 6 treatments: Anti-coagulants (Heparin), NSAIDs, Azithromycin, HCQ, Zinc replacement, and Glucocorticoids. As shown in Figure S3, all 6 treatments are associated with diminished effectiveness with increasing patient age, which is associated with severe cases of Covid-19 even after correcting for comorbidities, lab tests, and pre-admission treatments. Secondly, as shown in figure 5, NLR, a marker of inflammation and severe Covid-19 [14, 15], is associated with HTEs for 3 of these treatments. The statistical benefit of anti-coagulants, NSAIDs, and Azithromycin decrease with increased NLR, while the effectiveness of HCQ, Zinc, and GCs do not show statistically significant heterogeneity. We examine the association between GCs, NLR, and mortality in detail in the discussion.

**Figure 5:**
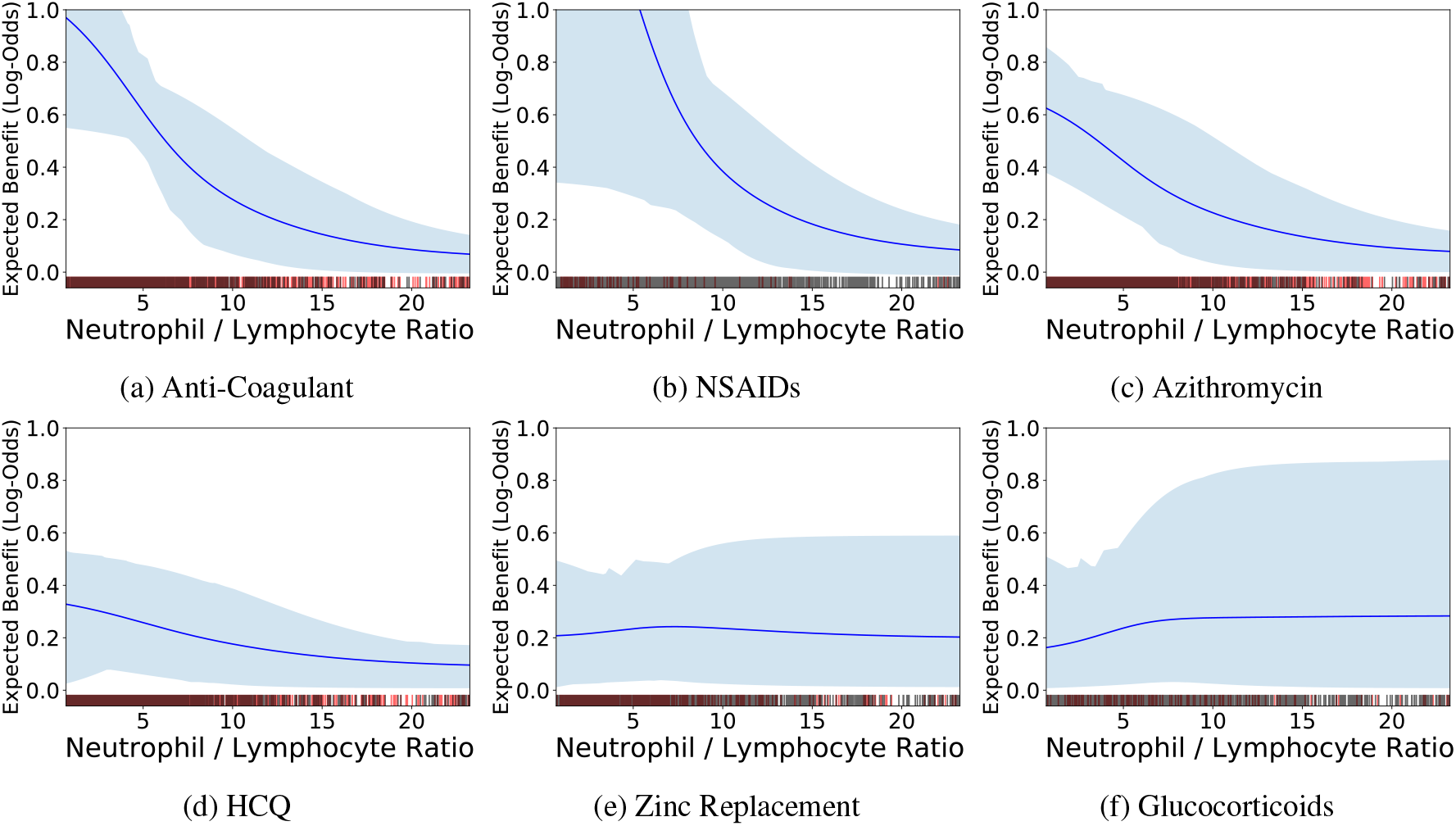
HTEs with respect to NLR. In each pane, we plot the estimated treatment benefit as a function of patient NLR, with shaded regions indicating 95% bootstrap confidence intervals. Red tick marks along the horizontal axis denote treated patients, while black tick marks denote untreated patients. NLR is a biomarker of inflammation and severe Covid-19; most of these treatments have lower effectiveness for higher NLR values. In contrast, zinc replacement and GCs appear to have to have stable or increasing effectiveness for larger values of NLR, consistent with an anti-inflammatory mechanism of action.

Next, we examine treatment-specific HTEs associated with comorbidities and concurrent treatments. These HTEs are summarized in Table 1; for each effect, we report the increased benefit associated with the interactive factor: 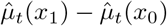 where *X*_1_ indicates that the interactive factor is present while *X*_0_ indicates that the interactive factor is not present. All of these benefit increases should be considered as an additive effect in coordination with the homogeneous treatment effects summarized in Figure 4.

**Table 1:** Heterogeneous Treatments Effects. We report all interactions in which the 95% confidence interval does not overlap a zero effect.

| Treatment | Interactive Factor | Benefit Increase (95% CI) |
| --- | --- | --- |
| GCs | Azithromycin | -0.28 (-0.54, -0.02) |
| NSAIDs | History of Alcoholism | 0.29 (0.05, 0.53) |
| Zinc Replacement | Congestive heart failure | -0.26 (-0.47, -0.05) |
| Anti-Coagulant | Congestive heart failure | -0.17 (-0.32, -0.02) |
|  | History of Hallucinogen Usage | 0.44 (0.09, 0.80) |
|  | History of Alcoholism | 0.82 (0.22, 1.43) |
| Azithromycin | History of Opioid Usage | -0.50 (-0.98, -0.03) |
|  | Congestive heart failure | -0.15 (-0.26, -0.04) |
|  | HCQ | 0.10 (0.02, 0.18) |
| HCQ | Congestive heart failure | -0.14 (-0.25, -0.03) |
|  | Myocardial infarction | -0.09 (-0.17, -0.01) |
|  | History of Alcoholism | 0.17 (0.01, 0.33) |
|  | Azithromycin | 0.24 (0.06, 0.42) |
|  | History of Hallucinogen Usage | 0.39 (0.04, 0.73) |

These results suggest GCs are associated with decreased benefit in combination therapy with Azithromycin. Next, NSAIDs are associated with increased benefit for patients with a history of alcoholism, a pro-inflammatory condition [17]. Zinc replacement is associated with diminished benefit for patients with congestive heart failure; this result would be expected if any protective effects of zinc were due to a mediation of thromboses [18] because patients with congestive heart failure are routinely prescribed anticoagulants and platelet aggregation inhibitors as out-patient medications. Similarly, anti-coagulants, which have been shown to have prevent mortality in Covid-19 patients with a likelihood of thromboses [19], appear to be less protective for patients with congestive heart failure, while increasing in effectiveness for patients with histories of substance abuse. Azithromycin is associated with decreased benefit for patients with history of opioid usage and congestive failure, with a mildly positive interaction with HCQ. Finally, HCQ is associated with beneift increase in patients who are at reduced risk of negative side effects including arrythmias (e.g., patients without a history of afibrillation or congestive heart failure); in addition, HCQ is associated with increased benefit in combination with Azithromycin and for patients with hallucinogen usage. These mild HTEs contrast against the lack of statistical evidence for any homogeneous treatment effectiveness for either Azithromycin or HCQ (Figure 4), suggesting further study on targeted and combination therapies may be needed to understand any potential benefits of these treatments for Covid-19.

## 4 Discussion

This method of estimating personalized effectiveness of multiple treatments is best considered as a tool for hypothesis generation to be followed up by targeted statistical tests. Here we examine a case study of manual exploration of the effectiveness of GCs modulated by NLR, which was suggested by the heterogeneous treatment effect model.

### Case Study: NLR and GCs

GCs have been shown to improve outcomes of patients with severe cases of Covid-19 [20]. However, criteria for prescribing GCs are currently limited. GC prescription is highly correlated with later admission dates and mortality is lower at later dates for a number of reasons. To correct for this confounding, as well as confounding from patient characteristics, we use the mortality risk model to correct for all risk which can be attributed to factors other than GCs. A full discussion of methodology is provided in Sec. S3.

Corresponding sample sizes and ARRs for the adjusted risk ratio (ARR)s, stratified by patient NLR value into three groups, are given in Table 2. Of these three ARRs, we observe statistically significant evidence of GC effects only for patients with NLR 6-25. We hypothesize that for patients who are not at risk of severe inflammation, GCs have little effect; for patients who are admitted with extremely high NLR, GCs may be insufficient. Finally, while elevated NLR is a strong predictor of mortality, it is not the only risk factor (Figure S1). These results suggest that GCs may have limited benefit to patients who are at high risk without having an elevated NLR. We encourage further investigation into treatments which could be effective for this set of patients.

**Table 2:** ARRs for patients treated with GCs compared to patients not treated with GCs, calculated by patient NLR. ARRs smaller than 1 indicate reduced mortality for patients treated with GCs, i.e., a beneficial effect. P-values are calculated by bootstrap resampling of the test set.

| NLR Values | N GC | N Control | ARR | 95% CI | P-Value (Uncorr.) |
| --- | --- | --- | --- | --- | --- |
| 0-6 | 79 | 1728 | 0.99 | 0.68-1.31 | 0.975 |
| 6-25 | 99 | 1079 | 0.74 | 0.49-0.99 | **0.042** |
| >25 | 15 | 116 | 0.90 | 0.58-1.22 | 0.549 |

#### Limitations

As with all analyses of observational data, this approach has several limitations. Firstly, while the machine learning optimization seeks to allocate effect sizes to most statistically reliable indicators, we do not use any side information (such as treatment mechanism of action or time-series data) to perform causal inference. In addition, in this study we have considered only binary indicators for each treatment, choosing to assume that providers are following dosage protocols to standardize care. Finally, while additive models are interpretable and accurate, they are still susceptible to statistical biases [21] which may cause different model classes to recover different effects from a single dataset. Further works should investigate the potential for other classes of additive models to corroborate or dispute these findings.

## 5 Conclusions

In this paper, we have proposed a method to estimate heterogeneous (varying) effectiveness of medical treatments by training additive models to estimate personalized treatment benefits and share statistical power between many treatments. We applied this method to mortality risk models of Covid-19 patients and uncovered evidence supporting two pathways of mortality: inflammation and thrombosis. We see that many treatments appear to have heterogeneous effectiveness; in particular, anti-inflammation treatments tend to be more effective for patients with lower likelihood of thromboses, while anti-coagulation treatments tend to be more effective for patients with lower likelihood of inflammatory attacks. We also see some evidence consistent with super-additive effectiveness of combinatorial treatments.

## Data Availability

Data include medical medical records which cannot be made public.
MIMIC-IV data are available at

https://physionet.org/content/mimiciv/0.4/

## Footnotes

1 https://github.com/interpretml/interpret

