## Supplement for "Automated Interpretable Discovery of Heterogeneous Treatment Effectiveness: A Covid-19 Case Study"

#### S1 Dataset Construction

Our dataset consists of 11080 total hospitalized patients who have lab-confirmed cases of Covid-19. To filter out patients who were hospitalized for reasons other than Covid-19, we excluded patients who have indicators of (1) pregnancy: outpatient prenatal vitamins, in-patient oxytocics, folic acid preparations; or (2) scheduled surgery: urinary tract radiopaque diagnostics, laxatives, general anesthetics, antiemetic/antivertigo agents, or antiparasitics. We also require that the patients have recorded temperature, age, BMI, and Admission Day. Finally, we remove patients who died within six hours of admission.

To correct for patient risk confounding, we observe pre-admission features including demographics, comorbidities, outpatient medications, initial in-patient vitals, and initial in-patient lab tests. We exclude any measurement taken within 24 hours of the patient mortality. The 45 total features are listed below.

- Demographics/Vitals:
  - Age
  - Sex
  - BMI
  - Day
  - Temperature
- Comorbidities:
  - Myocardial Infarction
  - Congestive Heart Failure
  - Peripheral Vascular Disease
  - Cerebrovascular Disease
  - Dementia
  - Chronic Obstructive Pulmonary Disease
  - Peptic Ulcer Disease
  - Mild Liver Disease
  - Diabetes without chronic complications
  - Diabetes with chronic complications
  - Hemiplegia or paraplegia
  - Renal disease
  - Cancer (any malignancy)
  - Metastatic solid tumor
  - Charlson score
  - Hypertension
  - Atrial fibrillation
  - Valve Replacement
  - Rheumatoid Arthritis
- Outpatient Medications taken before hospitalization (limited to medication classes taken by at least 100 patients):
  - Antihyperglycemic, Biguanide Type
  - Laxatives And Cathartics

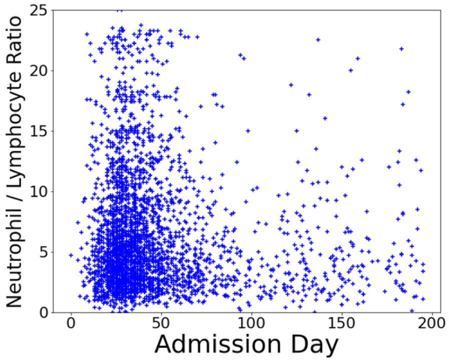


(a): Distribution of patients by NLR and Admission Day. Each marker indicates a single patient, with the vertical axis indicating NLR value on the initial lab test and the horizontal axis indicating the day the patient was admitted.

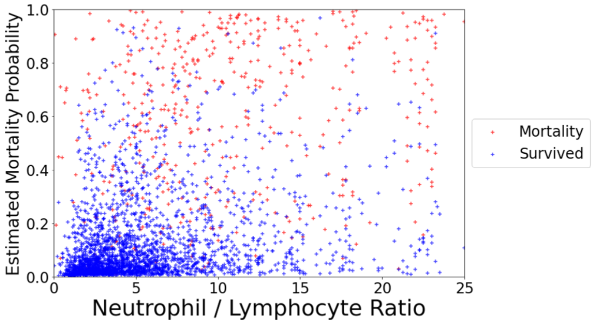


(b): Estimated probability of mortality for each patient, as predicted by the risk model operating on patient features at admission. While there is a relation between elevated NLR and increased risk of mortality, there are a sizable number of patients who have a large probability of mortality without elevated NLR.

Figure S1: Distribution of patients by admission day and NLR.

- Platelet Aggregation Inhibitors
- Vitamin D Preparations
- Calcium Channel Blocking Agents
- Proton-Pump Inhibitors
- Antihyperlipidemic-Hmgcoa Reductase Inhib (Statins)
- Beta-Adrenergic Agents, Inhaled, Short Acting
- Blood Sugar Diagnostics
- Anticonvulsants
- Analgesic/Antipyretics,Non-Salicylate
- Beta-Adrenergic Blocking Agents

• Lab Values:
  - Potassium (0.5% Missing)
  - Ferritin (8.3% Missing)
  - Calcium (0.5% Missing)
  - Neutrophil % (0.0% Missing)
  - Lymphocyte % (0.0% Missing)

Neutrophil-Lymphocyte Ratio is calculated by dividing Neutrophil % by Lymphocyte %. We exclude all patients who are missing either of these lab values.

The patient population has changed over time. The majority of patients were admitted from days 30-50. As Figure S1 shows, this period also contained the majority of patients with extremely elevated NLR levels. However, the range of NLR observed in the days and months following the initial peak of the pandemic remains wide. Many factors have shifted over the course of the pandemic, including mortality rates (Fig. S2a) and prescription patterns (Fig. S2b).

### S2 Heterogeneous Treatment Effectiveness with Respect to Patient Age

Figure S3 shows the expected benefit of several treatments with respect to patient age. The benefit of all treatments appears to decrease with respect to advanced patient age, however the confidence intervals are wide enough to allow a null effect for many of the treatments.

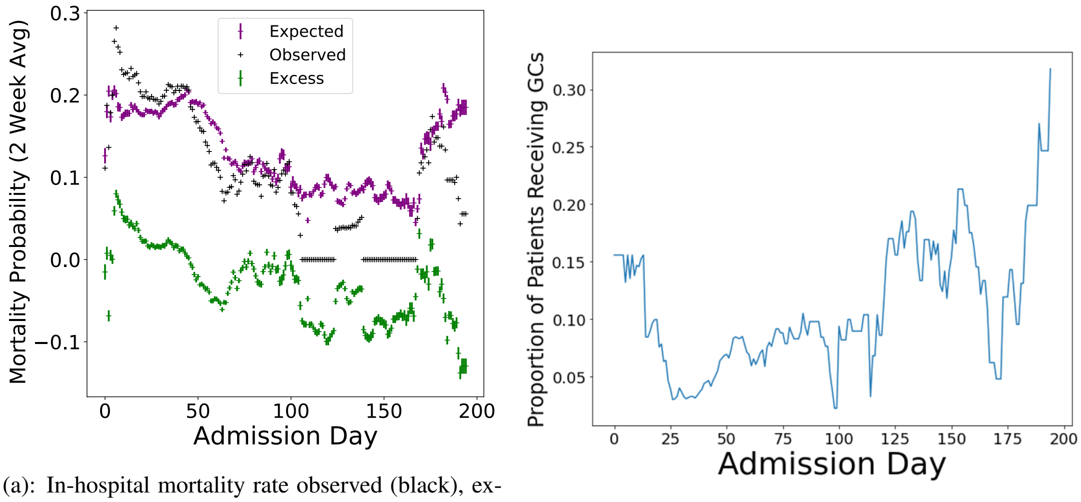


(a): In-hospital mortality rate observed (black), expected based on patient risk factors (purple), and the excess difference (green).

(b): The proportion of patients being prescribed glucocorticoids (GCs) has increased over time.

Figure S2: Mortality rate (left) and GC prescription (right) of hospitalized patients has changed over the course of the pandemic. The values in both plots are smoothed over a 14-day period by a running average.

### S3 Case Study: GCs and NLR

GCs have been shown to improve outcomes of patients with severe cases of Covid-19 [20]. However, criteria for prescribing GCs are currently limited. GC prescription is highly correlated with later admission dates and mortality is lower at later dates for a number of reasons. To correct for this confounding, as well as confounding from patient characteristics, we use the mortality risk model to correct for all risk which can be attributed to factors other than GCs. We are interested in

$$P_1(x, y) = P(\text{mortality}|X = x, \text{NLR} = y, \text{GC} = 1),$$
$$P_0(x, y) = P(\text{mortality}|X = x, \text{NLR} = y, \text{GC} = 0).$$

Comparing $P_1$ and $P_0$ separates mortality risk that can only be assigned to GC treatment out from mortality risk which could be assigned to other risk factors. The adjusted risk ratio (ARR) is the ratio $\frac{P_1}{P_0}$, while the adjusted risk difference (ARD) is the difference $P_1 - P_0$ [22, 23]. For both ARR and ARD, lower values indicate protective effects of GCs.

We follow the convention of [23] to estimate these quantities using a classification model. We use the additive model assumption that $P(\text{mortality}|X = x, \text{NLR} = y, \text{GC} = z) = f(x) + g(y, z)$. This means that a single $f$ (the background risk EBM) is shared between both groups of patients and that $g$ is an adjustment to this background risk.

For a small set of NLR values, we could estimate each $g(y_i, z_j)$ as the residual of the predictions of the background risk model on a set of held-out patients with NLR$= y_i$ and GC$= z_j$. However, for the continuous-valued NLR, we need a model to provide a smooth estimate of g. For this, we use a tree-based interaction available in `Interpret`. This whole procedure can be run as a native operation for the EBM model in `Interpret`, which trains and freezes main effects before estimating interaction effects. For robustness, we estimate $g$ on a test set of held-out patients which is resampled in each bootstrap iteration.

We first use this procedure to estimate the effect of GCs as a homogeneous benefit to all Covid-19 patients. We do not observe a strong benefit: mean ARR 0.96 (95% CI 0.86-1.09). Thus, we examine possible HTEs of GCs with benefit modulated by NLR level. Due to the limited sample size for patients with NLR>25, we focus our analysis on patients with NLR<25. Figures S4b and S4c show that the benefit of GCs is best for patients with NLR above 6 and the effect is largest in the region [6, 10].

Estimating HTEs for all NLR values greatly reduces statistical power and inflates the width of the CIs. To test for statistical significance of the GC benefit, we group NLR values into 3 ranges: NLR < 6, NLR 6-25, NLR >25.

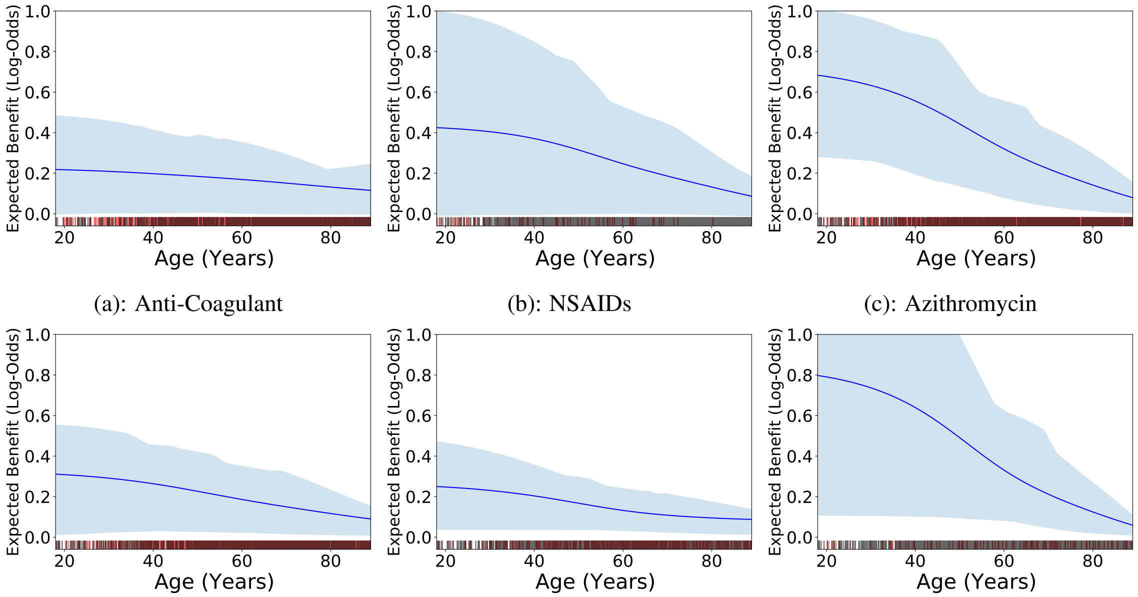


(a): Anti-Coagulant (b): NSAIDs (c): Azithromycin

(d): HCQ (e): Zinc Replacement (f): Glucocorticoids

Figure S3: The effectiveness of all medications appears to decrease with increased patient age.

Corresponding sample sizes and ARRs are given in Table 2. Of these three ARRs, we observe statistically significant evidence of GC effects only for patients with NLR 6-25. We hypothesize that for patients who are not at risk of severe inflammation, GCs have little effect; for patients who are admitted with extremely high NLR, GCs may be insufficient.

Finally, while elevated NLR is a strong predictor of mortality, it is not the only risk factor (Figure S1). Our results suggest that GCs may have limited benefit to patients who are at high risk without having an elevated NLR. We encourage further investigation into treatments which could be effective for this set of patients.

### S4 Method Validation with MIMIC-IV

To validate this new methodology for hypothesis generation of multiple heterogeneous treatment effects, we apply the method to a large, recent dataset of ICU patients. The Medical Information Mart for Intensive Care (MIMIC)-IV database [24, 25] provides deidentified critical care data for over 50,000 patients admitted to intensive care units (ICU) at the Beth Israel Deaconess Medical Center (BIDMC). In this analysis, we use all 53,150 patients admitted to the ICU in MIMIC-IV, spanning 69,211 hospital admission events and 76,540 ICU stays. Our outcome is in-hospital mortality, and we use demographics, comorbidities, and in-hospital medications analogous to the procedure used for the Covid-19 dataset. All heterogeneous treatment effects with change in mortality log-odds greater than 0.1 are provided in Figures S5-S8.

In short, our method identifies that nearly all treatments are more effective at preventing mortality for younger patients than for older patients, after correcting for all other factors. There are however, several notable exceptions to this trend. The effectiveness of Dextrose 5% increases for increasing age (Fig. S5e), in concordance with a diminished control of blood sugar in older patients. In addition, the effectiveness of dialysis increases with increasing age (Fig. S5f), in concordance with increased attention and motivation required for the routine necessary for dialysis. Finally, the effectiveness of fresh frozen plasma (Fig. S5j) and packed red blood cells (Fig. S7d) both increase with increasing age, in concordance with a diminished ability for elderly patients to synthesize blood cells and an increased reliance on transfusions.

In addition to age, several treatments have effectiveness which varies with respect to the patient's hemoglobin concentration. In particular, prophylactic heparin (Fig. S8a) and Potassium Chloride (Fig. S8b) appear to have increased

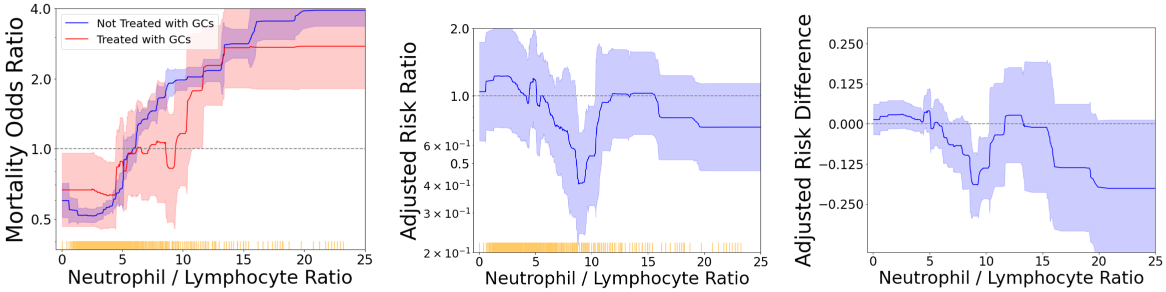


(a): Mortality Risk (b): ARR (c): ARD

Figure S4: (a) Mortality risk conferred by NLR in addition to all other risk factors, with patients stratified by glucocorticoid treatment. We plot the risk curve estimated by the generalized additive model, with 95% confidence intervals shaded. Each yellow tick mark along the horizontal axis indicates 10 patients. (b,c) Estimated benefit of glucocorticoid treatment after correcting for patient risk, stratified by NLR value. Shaded regions indicate 95% CIs. Despite the small sample size in each NLR bin, there is still statistical significance near NLR=8.5.

effectiveness for increased hemoglobin. Heparin is an anti-coagulant, which is most beneficial for patients at risk for thromboses; Potassium Chloride causes Chloride shift which reduces the oxygen affinity of hemoglobin [26].

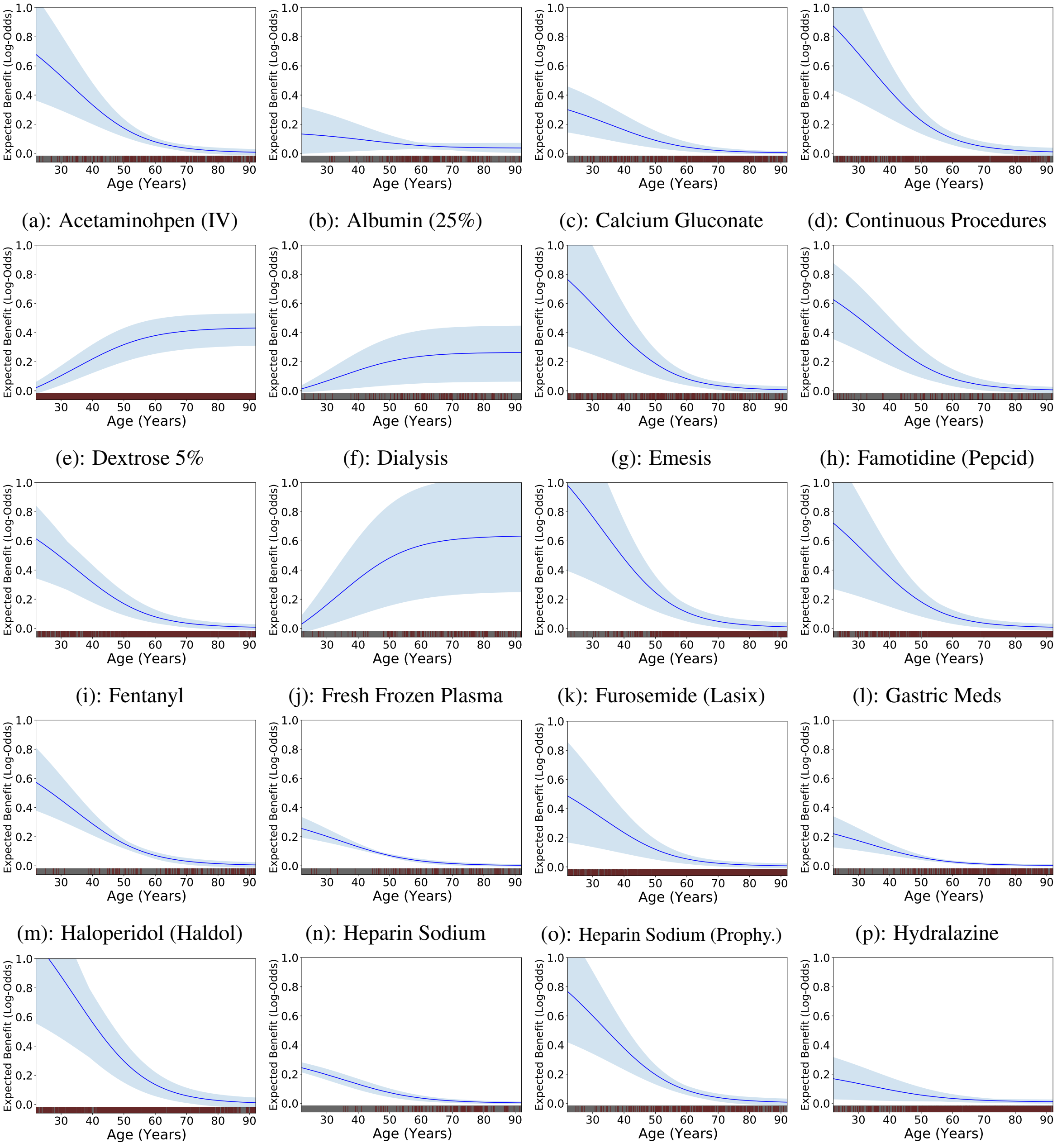


(a): Acetaminohpen (IV) (b): Albumin (25%) (c): Calcium Gluconate (d): Continuous Procedures
(e): Dextrose 5% (f): Dialysis (g): Emesis (h): Famotidine (Pepcid)
(i): Fentanyl (j): Fresh Frozen Plasma (k): Furosemide (Lasix) (l): Gastric Meds
(m): Haloperidol (Haldol) (n): Heparin Sodium (o): Heparin Sodium (Prophy.) (p): Hydralazine
(q): Hydromorphone (Dilaudid) (r): Ileoconduit (s): Insulin Glargine (t): Insulin Humalog

Figure S5: Age-Dependent Treatment Effectiveness in MIMIC-IV (cont.)

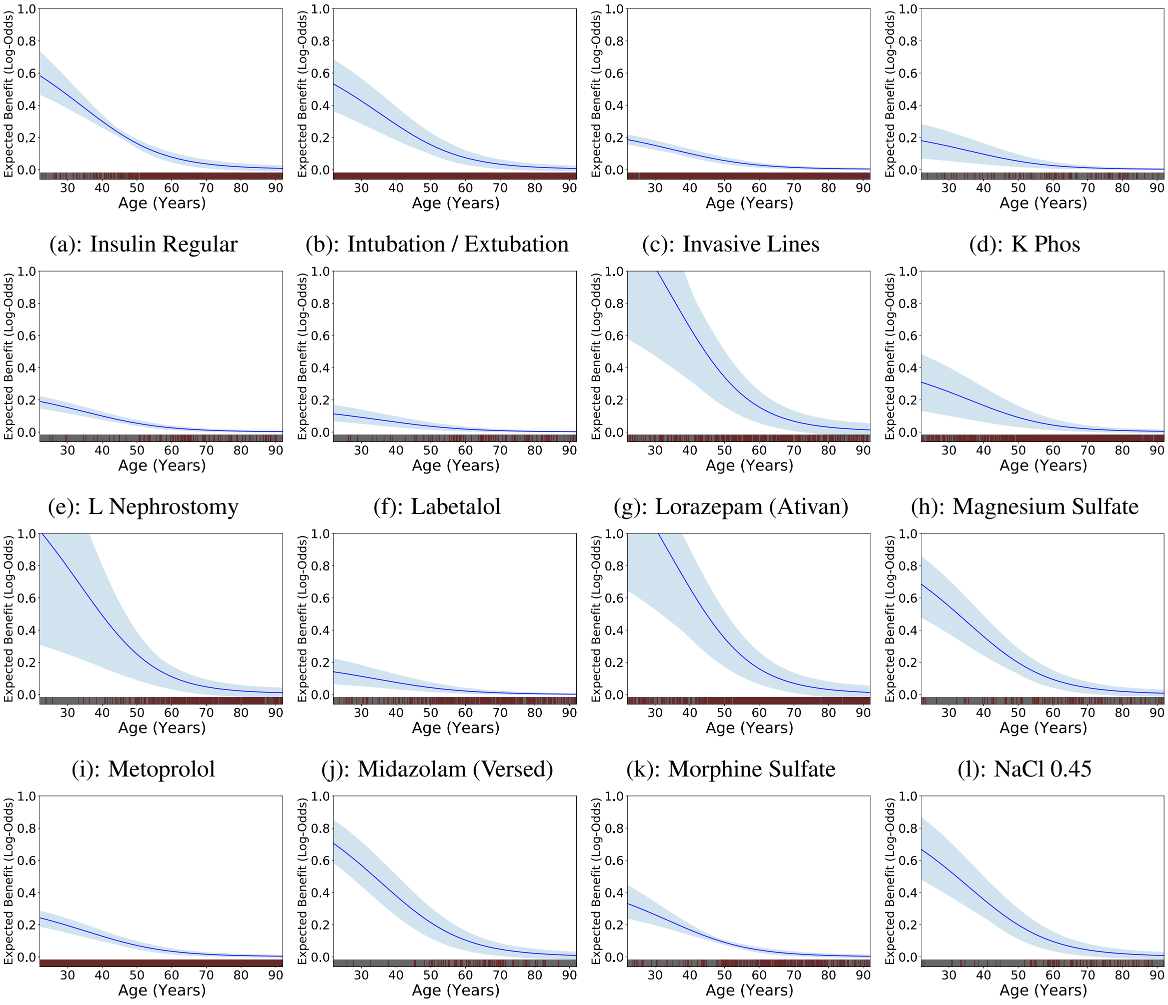


(a): Insulin Regular
(b): Intubation / Extubation
(c): Invasive Lines
(d): K Phos
(e): L Nephrostomy
(f): Labetalol
(g): Lorazepam (Ativan)
(h): Magnesium Sulfate
(i): Metoprolol
(j): Midazolam (Versed)
(k): Morphine Sulfate
(l): NaCl 0.45
(m): NaCl 0.9
(n): OR Cell Saver Intake
(o): OR Crystalloid Intake
(p): Omeprazole (Prilosec)

Figure S6: Age-Dependent Treatment Effectiveness in MIMIC-IV (cont.)

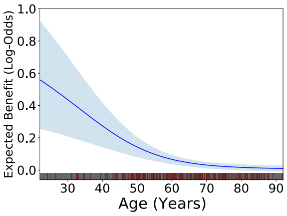


(a): Ostomy (output)

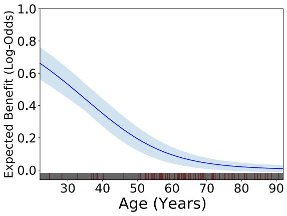


(b): PACU Crystalloid Intake

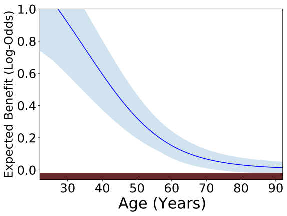


(c): PO Intake

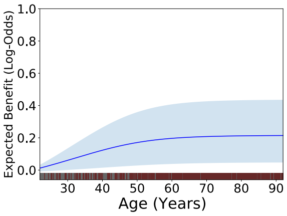


(d): Packed Red Blood Cells

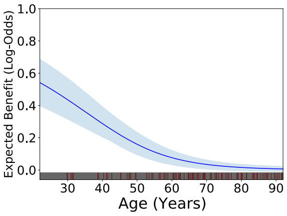


(e): Pantoprazole (Protonix)

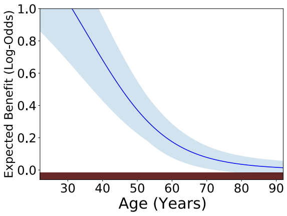


(f): Peripheral Lines

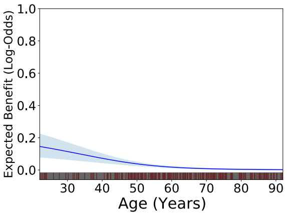


(g): Piggyback

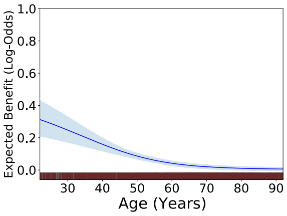


(h): Potassium Chloride

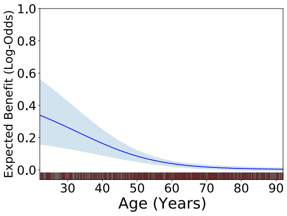


(i): Propofol

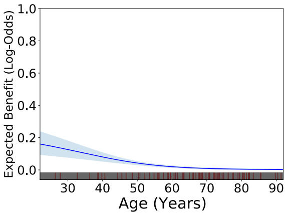


(j): R-Pleural 1

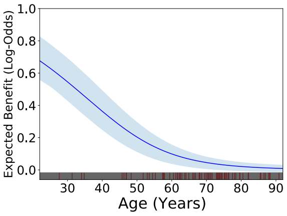


(k): Rantidine (Prophylaxis)

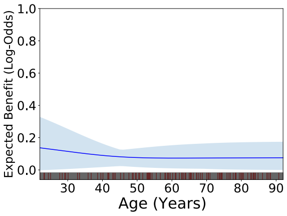


(l): Sodium Bicarbonate 8.4

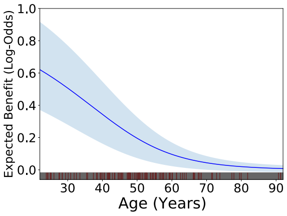


(m): Thiamine

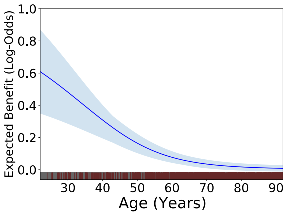


(n): Tubes

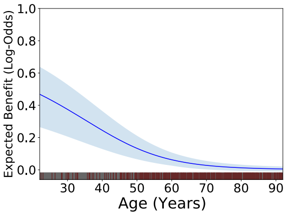


(o): Ventilation

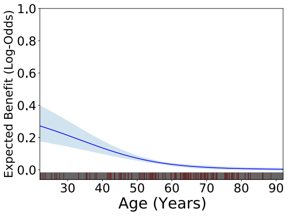


(p): Wound Vac

Figure S7: Age-Dependent Treatment Effectiveness in MIMIC-IV (cont.)

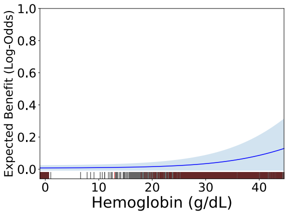


(a): Heparin Sodium (Prophy.)

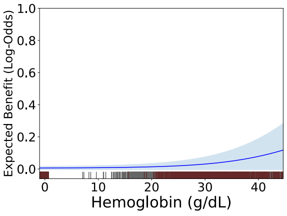


(b): Potassium Chloride

Figure S8: Hemoglobin-Depedent Treatment Effectiveness in MIMIC-IV
